# Emergence of a SARS-CoV-2 E484K variant of interest in Arizona

**DOI:** 10.1101/2021.03.26.21254367

**Authors:** Peter T. Skidmore, Emily A. Kaelin, LaRinda A. Holland, Rabia Maqsood, Lily I. Wu, Nicholas J. Mellor, Joy M. Blain, Valerie Harris, Joshua LaBaer, Vel Murugan, Efrem S. Lim

## Abstract

SARS-CoV-2 is locked in a high-stakes arms race between the dynamics of rising population immunity and escape mutations. The E484K mutation in the spike protein reduces neutralization by post-vaccination sera and monoclonal antibody therapeutics. We detected the emergence of an E484K harboring variant B.1.243.1 from a common circulating variant (B.1.243) in the United States. In contrast to other instances when the E484K mutation was acquired independently in the parental lineage, genomic surveillance indicates that the B.1.243.1 variant of interest is in the process of being established in Arizona and beginning to cross state borders to New Mexico and Texas. Genomic, epidemiologic and phylogenetic evidence indicates that the B.1.243.1 variant of interest is poised to emerge. These findings demonstrate the critical need to continue tracking SARS-CoV-2 in real-time to inform public health strategies, diagnostics, medical countermeasures and vaccines.

## Main

The development of multiple effective vaccines is a major advancement in the fight against COVID-19. However, SARS-CoV-2 continues to evolve mutations in its genome as it spreads around the world. Although many mutations have little or no consequence on virus fitness, some mutations affect receptor binding, reduce antibody neutralization or affect diagnostic tests^1-4^. Other mutations have been associated with increased transmission and clinical disease severity^5,6^. Based on the combination of mutations and associated attributes, variants can be elevated to the classification level of variant of interest (VOI), variant of concern (VOC) and variant of high consequence (VOHC)^7^. Current VOCs include B.1.1.7 that emerged from the UK, B.1.351 that was first identified in South Africa^8^, P.1 that was first described in Brazil^9^, and B.1.427 and B.1.429 that were first detected in California in the US^10^. Additionally, there are 3 VOIs – all of which harbor the E484K mutation in the spike glycoprotein S gene: B.1.525 and B.1.526 both first detected in New York in the US^11^, and P.2 that was first identified in Brazil^12^.

In an effort to provide state-wide genomic surveillance, we sequenced the SARS-CoV-2 genome from 688 positive samples collected from December 28 2020 to March 13 2021 in Arizona, USA. 638 high-quality complete genomes (92.7%) were successfully sequenced that included variants such as B.1.1.7 (n=49, 7.7%), B.1.427/429 (n=214, 33.5%) and P.2 (n=6, 0.9%). We detected 7 genomes associated with a common B.1.243 variant that had acquired an E484K mutation in the spike protein. The novel variant had 11 lineage-defining mutations including V213G and E484K in the spike gene, a 9-nt deletion in ORF1ab (ΔSGF3675-77), a 3-nt insertion in the non-coding intergenic region upstream of the N gene and other synonymous substitutions (**Figure 1A, Supplementary Table 1**). These 11 conserved mutations are distinct from the mutations associated with the parental lineage, B.1.243. The parental B.1.243 lineage is a common circulating variant in the US that was first observed at the start of the pandemic as early as March 2020 (**Figure 1B**, 96.9%). The B.1.243 parent lineage encodes the spike gene D614G substitution, but none of the other concerning mutations (**Figure 1A, Supplementary Table 2**). Therefore, we designate the new E484K harboring variant the provisional name of B.1.243.1.

**Figure 1.**
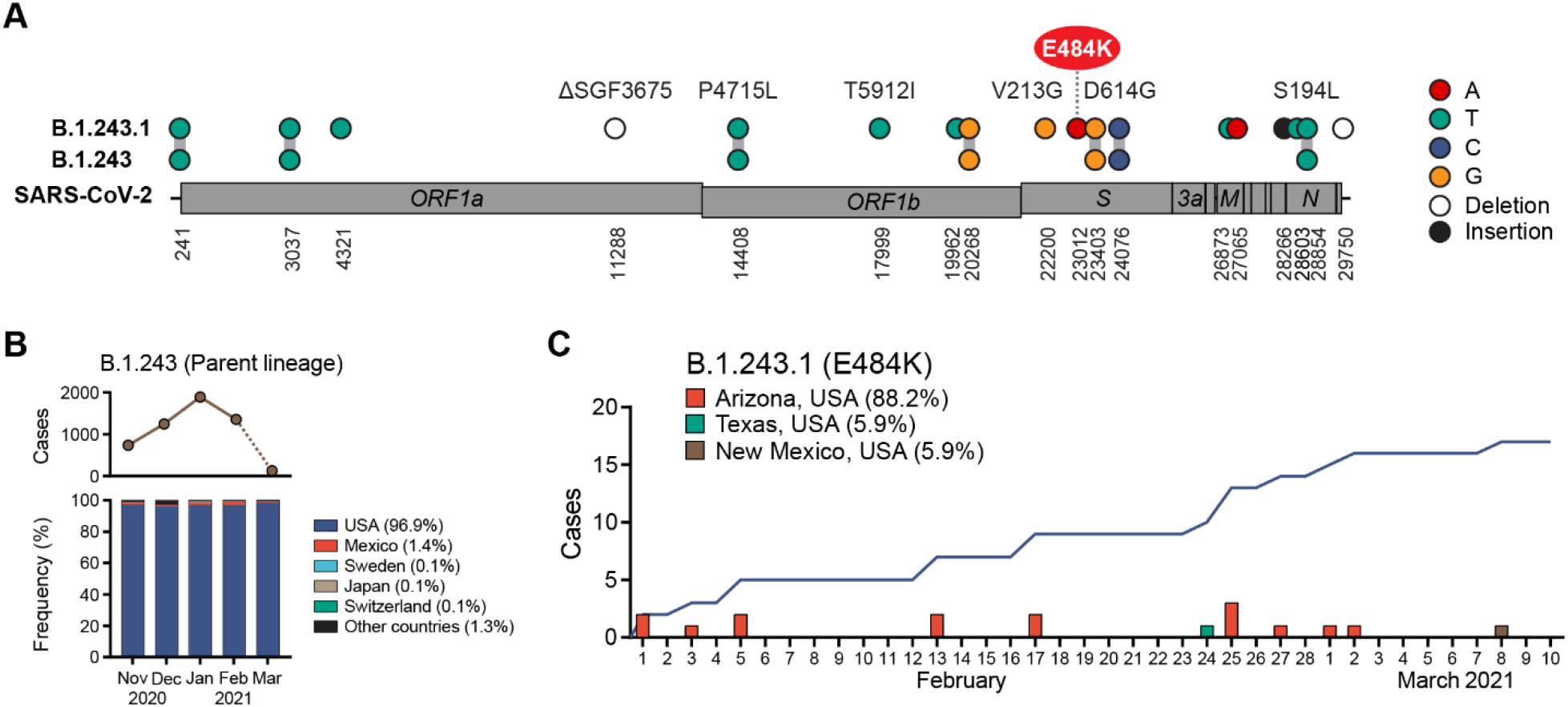
Emergence of E484K harboring B.1.243.1 variant in Arizona, USA. **(A)** B.1.243.1 lineage-defining mutations are shown on the SARS-CoV-2 genome. Mutations are shown in reference to the SARS-CoV-2 Wuhan-1 genome position (NC_045512.2). **(B)** Global prevalence of B.1.243 parental lineage from November 2021 to March 2021. Monthly number of B.1.243 cases (top) and distribution by country (bottom) are shown. Average frequency for each country is shown in parenthesis. Sequences from March are under-reported at the time of this reporting (indicated by dashed line). **(C)** B.1.243.1 case incidence reporting from February to March 2021. Cumulative case incidence is plotted as a line graph.

We next examined the GISAID repository for additional B.1.243.1 genomes to understand its prevalence and geographic distribution. We found that B.1.243.1 is predominantly established in Arizona. Of the 17 cases of B.1.243.1 sequenced to date, 15 cases were from Arizona ranging from February 1 to the most recent case on March 2 (**Figure 1C, Supplementary Table 3**). 1 case was sequenced from a sample collected in Houston, Texas on February 24 and the most recent case from a sample collected in New Mexico on March 8, suggesting that B.1.243.1 has spread to other states in the US. We also identified 2 instances where the parental B.1.243 lineage independently acquired the E484K mutation. However, in contrast to the new B.1.243.1 variant, both genomes lacked the 11 B.1.243.1 lineage-defining mutations and appear to be dead-end transmission events. Phylogenetic analyses support that the B.1.243.1 sequences form a monophyletic clade within the 20A/B.1.243 clade (**Figure 2**). Multiple internal branching observed in the B.1.243.1 clade indicate continued diversification of the lineage sequences. This suggests that B.1.243.1 is being established in circulation within Arizona. In contrast, the two B.1.243 cases bearing the E484K mutation alone were phylogenetically distinct from the B.1.243.1 clade, supporting that they had evolved independently.

**Figure 2.**
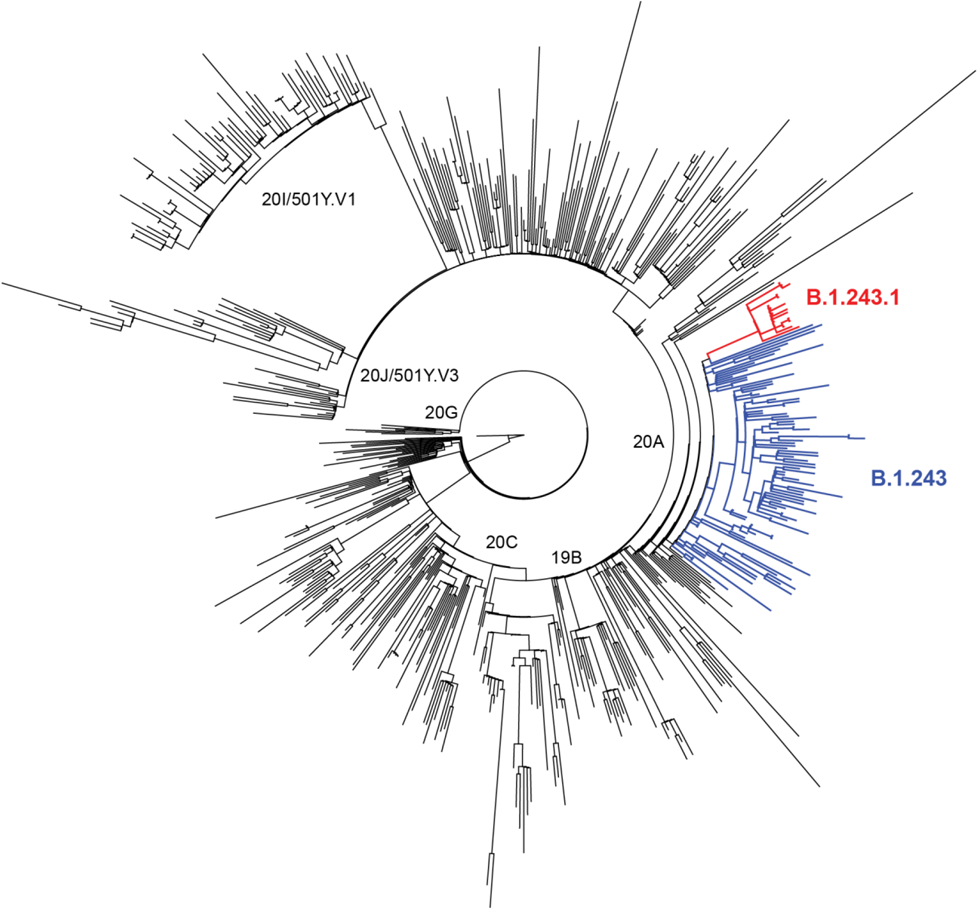
Phylogenetic relationship of the novel B.1.243.1 lineage. Maximum likelihood phylogeny of diverse SARS-CoV-2 sequences including 500 representative global sequences, 100 B.1.243 parent lineage sequences, and the 17 B.1.243.1 sequences. The novel B.1.243.1 lineage is indicated in red branches (clade bootstrap support: 100), and the parental B.1.241 lineage in blue.

Mutations in the SARS-CoV-2 spike protein receptor binding domain (RBD) can affect ACE2 receptor binding and antibody neutralization^3,13,14^. E484K variants have reduced neutralization by monoclonal antibodies and post-vaccination sera^1,13,15-17^, potentially complicating antibody-based countermeasures and vaccines. E484K variants have also been identified in reinfection cases, suggesting a role in breakthrough infections^12,18^. The impact of E484K mutation on spike-ACE2 interaction and binding affinity is less clear^19-21^. Little is known about the functional consequences of the other B.1.243.1 lineage-defining mutations and whether they contribute to the successful transmission in Arizona. Human behavioral risk factors and public policies (e.g. reopening) can also impact the transmissibility of SARS-CoV-2^22,23^. Due to limited sequencing efforts in the US, the true prevalence and geographic distribution of B.1.243.1 is not well understood. However, this apparent limitation can be resolved with increased sequencing surveillance together with contact tracing. Another limitation of this study was that only 92.7% of samples were successfully sequenced. Most of the partial genomes sequenced were from samples that had lower viral load (high CT values in the 30s), which further reduced the number of sequences that could be used to analyze for this variant.

Based on the mutation profile, regional introduction and phylogenetic evidence, we recommend vigilant monitoring of B.1.243.1 as a potential variant of interest. This study demonstrates the need for sustained genomic surveillance in public health strategies and responses.

## Materials and Methods

### Study population

As part of SARS-CoV-2 genomic surveillance efforts, saliva samples submitted for COVID-19 testing to the Arizona State University’s Biodesign Clinical Testing Laboratory (ABCTL) that tested positive (TaqPath COVID-19 Combo Kit, Applied Biosystems) were randomly selected for next-generation sequencing^24^. Samples covered a broad distribution from counties across Arizona, United States. This study was approved by the Arizona State University Institutional Review Board.

### SARS-CoV-2 sequencing

RNA was extracted from 250 µl of saliva sample (KingFisher Flex, Thermo Scientific) according to the manufacturer’s guidelines. First-strand cDNA synthesis was performed using random hexamers (SuperScript III/IV Reverse Transcriptase, Life Technologies), followed by PCR amplification of tiled amplicons spanning the SARS-CoV-2 genome (Swift Normalase Amplicon Panel, Swift Biosciences) and library construction. Libraries were sequenced on the Illumina MiSeq (version 2, 2×150) and NextSeq 500 (version 2.5, 2×150, mid or high output).

### Sequencing analysis

Illumina sequencing reads were quality filtered to remove adaptors and low-quality bases using BBTools. High-quality-filtered reads were mapped to the SARS-CoV-2 Wuhan1 reference genome (NC_045512.2) using BWA-MEM^25^ and amplicon primers were trimmed using Primerclip (version 0.3.8)^26^. Consensus sequence was called using iVar (version 1.0; parameters -q 20, -t 0.75, -m 20, -n N)^27^. Sequence alignments were performed with MAFFT (version 7.471)^28^ and variant calling using Geneious Prime (version 2021). Sequences used in phylogenetic analysis include the global sequences from GISAID^29^ subset to 500 sequences, a subset of 100 B.1.243 sequences from all B.1.243 GISAID sequences, and the 17 B.1.243.1 lineage sequences. Phylogenetic reconstruction was performed with IQTree (version 2.0.3^30^, iqtree -nt AUTO -bb 1000 -m MFP -mset GTR), and Augur (version 11.3.0)^31^.

## Data Availability

Sequence data have been deposited in GISAID.

## Data availability

Sequence data have been deposited in GISAID.

## Acknowledgements

We gratefully acknowledge the authors from originating laboratories responsible for obtaining the specimens and the submitting laboratories where genetic sequence data were generated and shared via the GISAID initiative, on which part of the research is based. This work was supported in part by the Arizona State University Knowledge Enterprise and Arizona Department of Health Services.

## Contributions

Methodology: P.T.S., E.A.K., L.A.H., R.M., L.I.W., N.J.M., J.M.B., V.H., E.S.L.; Investigation: P.T.S., E.A.K., R.M., E.S.L.; Resources: L.I.W., V.H., J.L., V.M.; Data curation: P.T.S., E.A.K., R.M.; Writing-original draft: P.T.S., E.A.K., R.M., E.S.L.; Writing-review and editing: P.T.S., E.A.K., L.A.H., R.M., E.S.L.; Supervision: J.L., V.M., E.S.L.; Conceptualization: E.S.L.; Funding acquisition: E.S.L. All authors reviewed and approved the final manuscript.

---

**Supplementary Table 1.** B.1.243.1 lineage-defining mutations.

| **Gene** | **Nucleotide change** | **Amino acid change** | **Number of B.1.243.1 genomes with mutation** |
| --- | --- | --- | --- |
| ORF1ab | C4321T | Synonymous | 17/17 |
| ORF1ab | 11288-11296 deletion | SGF 3675-3677 deletion | 17/17 |
| ORF1ab | C17999T | T5912I | 17/17 |
| ORF1ab | G19962T | Synonymous | 17/17 |
| S | T22200G | V213G | 17/17 |
| S | G23012A | E484K | 17/17 |
| M | C26873T | Synonymous | 17/17 |
| M | G27065A | Synonymous | 17/17 |
| Non-coding, upstream of N | 28266 GCC insertion | Non-coding | 17/17 |
| N | C28603T | Synonymous | 17/17 |
| 3' UTR | 29750-29761 deletion | Non-coding | 13/13* |

All genome positions are in reference to the SARS-CoV-2 Wuhan-1 sequence (NC\_045512.2).
\*4 sequences were omitted due to ambiguous (N) nucleotides.

**Supplementary Table 2.**
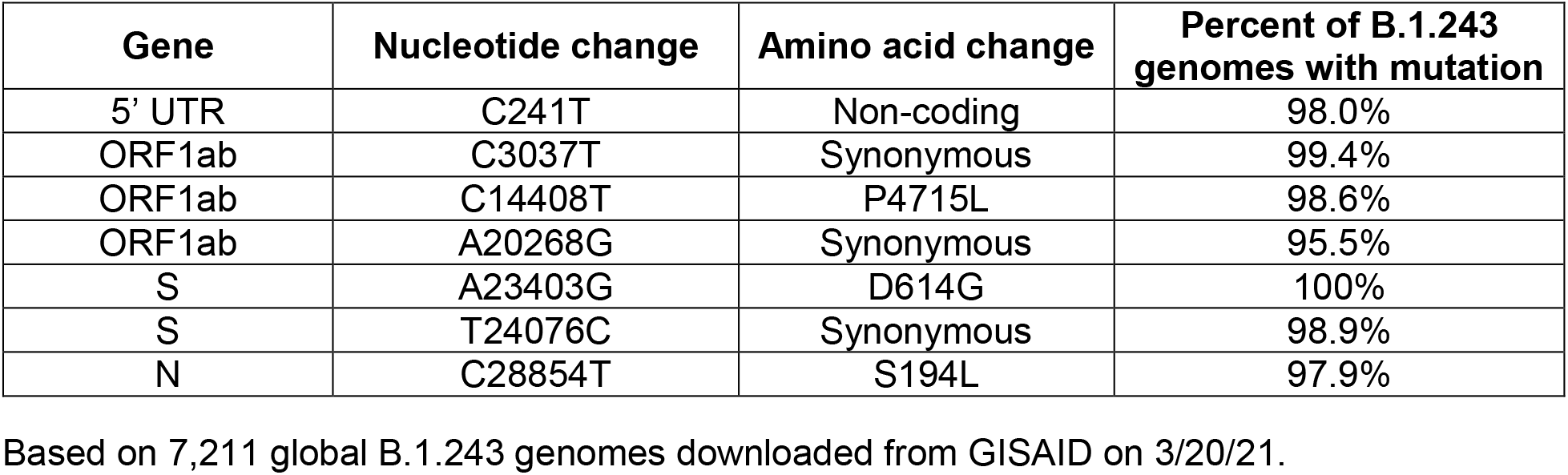
Lineage-defining mutations in parental B.1.243 lineage.

**Supplementary Table 3:** B.1.243.1 sequences.

| **Name** | **GISAID**<br>**Accession** | **Location** | **Collection**<br>**Date** |
| --- | --- | --- | --- |
| hCoV-19/USA/AZ-ASU2621/2021 | EPI_ISL_1364812 | Arizona | 2/1/2021 |
| hCoV-19/USA/AZ-ASU2625/2021 | EPI_ISL_1364644 | Arizona | 2/1/2021 |
| hCoV-19/USA/AZ-ASU2857/2021 | EPI_ISL_1364649 | Arizona | 2/3/2021 |
| hCoV-19/USA/AZ-CDC-21801839/2021 | EPI_ISL_1090700 | Arizona | 2/5/2021 |
| hCoV-19/USA/AZ-CDC-21802041/2021 | EPI_ISL_1090853 | Arizona | 2/5/2021 |
| hCoV-19/USA/AZ-CDC-22062741/2021 | EPI_ISL_1139766 | Arizona | 2/13/2021 |
| hCoV-19/USA/AZ-ASU2754/2021 | EPI_ISL_1364775 | Arizona | 2/13/2021 |
| hCoV-19/USA/AZ-ASU3132/2021 | EPI_ISL_1365543 | Arizona | 2/17/2021 |
| hCoV-19/USA/AZ-ASU2925/2021 | EPI_ISL_1365483 | Arizona | 2/17/2021 |
| hCoV-19/USA/TX-HMH-MCoV-29140/2021 | EPI_ISL_1303700 | Texas | 2/24/2021 |
| hCoV-19/USA/AZ-ASU2540/2021 | EPI_ISL_1291671 | Arizona | 2/25/2021 |
| hCoV-19/USA/AZ-TG758899/2021 | EPI_ISL_1292269 | Arizona | 2/25/2021 |
| hCoV-19/USA/AZ-TG758666/2021 | EPI_ISL_1292117 | Arizona | 2/25/2021 |
| hCoV-19/USA/AZ-CDC-22555310/2021 | EPI_ISL_1290985 | Arizona | 2/27/2021 |
| hCoV-19/USA/AZ-CDC-22554229/2021 | EPI_ISL_1290992 | Arizona | 3/1/2021 |
| hCoV-19/USA/AZ-TG761699/2021 | EPI_ISL_1296905 | Arizona | 3/2/2021 |
| hCoV-19/USA/NMDOH-2021075279/2021 | EPI_ISL_1340909 | New Mexico | 3/8/2021 |

